# Kidney Status and Events Preceding Death in Heart Failure: A Real-world Nationwide Study

**DOI:** 10.1101/2024.10.15.24315570

**Authors:** Deewa Zahir Anjum, Caroline Hartwell Garred, Nicholas Carlson, Emil Fosbol, Mariam Elmegaard, Pardeep S. Jhund, John J.V. McMurray, Mark C. Petrie, Lars Kober, Morten Schou

## Abstract

**Background:** Chronic kidney disease (CKD) is a frequent comorbidity in heart failure (HF) patients, affecting prognosis and mortality. This study investigates the relationship between kidney function and adverse kidney events preceding death in HF patients.

**Methods:** We analyzed registry data of HF patients who died between 2014 and 2021, with at least one year of HF diagnosis. Adverse kidney events, including acute kidney injury (AKI) and end-stage kidney disease (ESKD), were assessed. Patients were grouped by estimated glomerular filtration rate (eGFR) one year before death: eGFR≥60, eGFR 30-59, and eGFR<30.

**Results:** Among 36,435 HF patients who died, 37% had eGFR≥60, 46% had eGFR 30-59, and 17% had eGFR<30 one year before death. Median age was 81 years, and 61.2% were men. Adverse kidney events occurred in 13.1% of patients. AKI was inversely related to kidney function, affecting 6.5% (95% confidence interval 6.1-6.9) of those with eGFR≥60, 7.0% (6.6.-7.4) with eGFR 30-59, and 21.9% (20.9-22.9) with eGFR<30. ESKD occurred in 0.7%(0.6-0.9), 2.6% (2.4-2.8), and 35.5% (34.3-36.7) of patients in the respective eGFR categories. In the last three months before death, kidney function notably declined, with increased chronic kidney replacement therapy. Factors associated with higher adverse kidney events included younger age, male sex, in-hospital death, and greater frailty.

**Conclusions:** In HF patients, AKI and ESKD are common in the last year of life, particularly in those with lower baseline eGFR, with kidney decline accelerating in the final months.

## Background

Despite advances in HF management, patients with concurrent HF and kidney dysfunction are at higher risk of adverse events ^1^, including cardiovascular events^2^ and progression to end-stage kidney disease^3^. Kidney dysfunction accounts for approximately one million deaths globally each year ^4^, with an inverse relationship observed between estimated glomerular filtration rate (eGFR) and the risk of cardiovascular mortality^5^, particularly sudden death^6^. Individuals with severe kidney dysfunction (eGFR <25 mL/min/1.73 m²) are frequently excluded from HF clinical trials^7,8^.

While the renewed attention to kidney health has led to increasing inclusion of kidney-related outcomes in major HF trials^9–11^, specific data on kidney-related mortality, where kidney dysfunction is a critical factor, are still sparse^12^. Many HF patients experience an accelerated decline in eGFR over time^13^, however the incidence of and temporal relationship between adverse kidney events and death in patients with HF are not well documented. This study analyzes real-world data to explore the incidence of kidney events preceding death in HF patients, aiming to clarify patterns of kidney dysfunction patterns in the terminal phase of HF. The findings could guide early intervention strategies, improve prognosis, and influence clinical trial designs to better include those with severe kidney dysfunction, enhancing the the applicability of research findings.

## Methods

### Data sources

We obtained data from the nationwide registers in Denmark. All Danish citizens are enrolled in the Civil Registration Registry and assigned a unique personal number, which facilitates identification across multiple national registries^14^. We utilized data from the following three Danish national registries: (i) The Danish National Patient Registry, which records all hospital inpatient and outpatient contacts (coded using the 10th edition of the International Classification of Diseases (ICD-10) codes)^15^ and surgical procedures (coded according to the Nordic Medico-Statistical Committee Classification of Surgical Procedures), (ii) the Register of Laboratory Results for Research, which contains laboratory test results^16^, and (iii) The National Prescription Registry, which captures data on all medical prescriptions filled at Danish pharmacies using the Anatomical Therapeutic Chemical Classification System (ATC) codes^17^.

### Study population

We included all deaths that occurred between January 1, 2014, and December 31, 2021, in patients previously diagnosed with HF. Patients were included in the study if they died after at least one year with HF. We excluded individuals who had missing information on estimated glomerular filtration rate (eGFR). Baseline was defined as one year prior to death. The exposure assessment period correspondingly extended up to one year before death, except for kidney transplantation, which had a 15-year lookback period (as illustrated in Figure 1). Patients were categorized into eGFR groups based on their eGFR at baseline, measured either one year before death or up to 90 days before baseline: eGFR≥60 mL/min/1.73 m^2^, eGFR 30-59 mL/min/1.73 m^2^, and eGFR<30 mL/min/1.73 m^2^. If multiple eGFR values were available, the one closest to the baseline was used. The eGFR was calculated using the 2021 CKD-EPI creatinine formula^18^.

**Figure 1:**
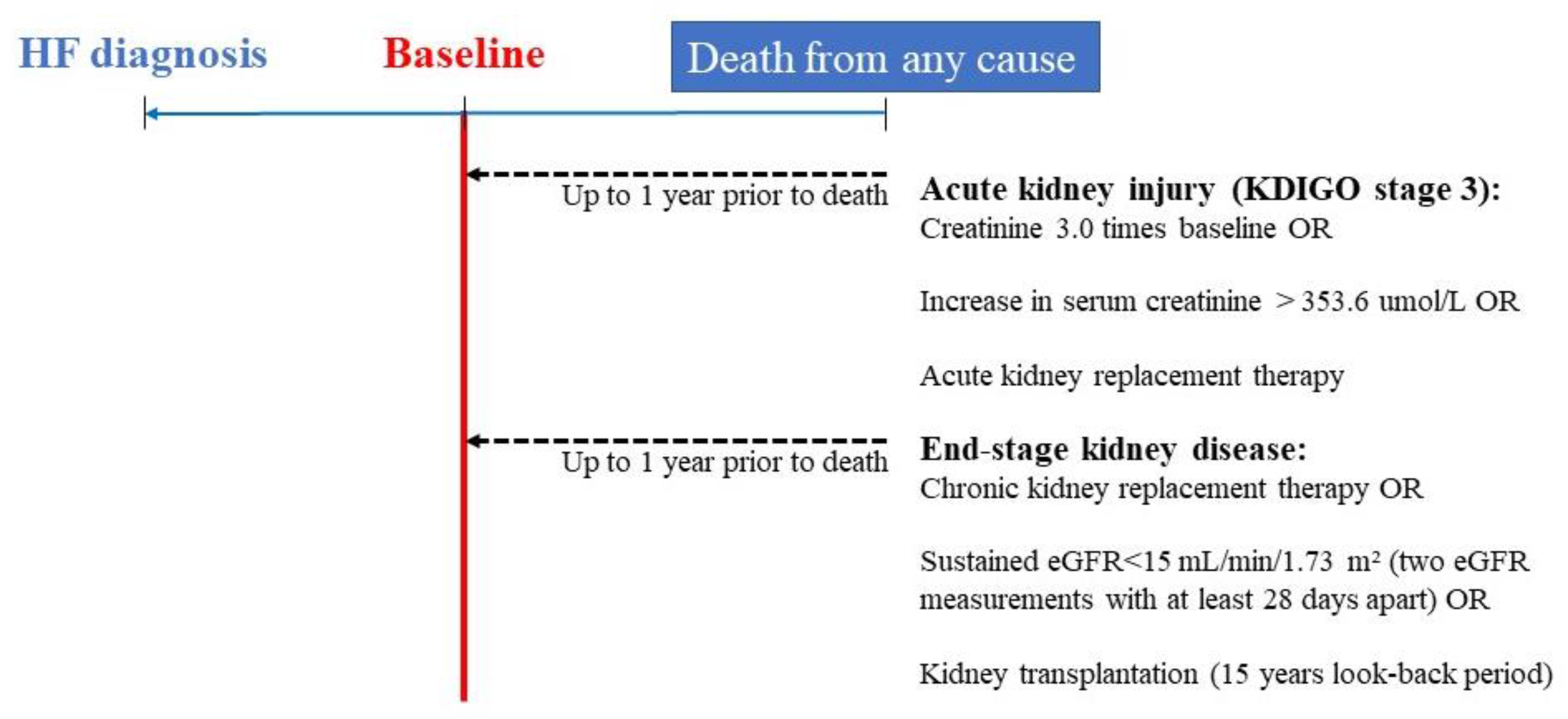
Study timeline

### Outcomes

The outcome of interest was adverse kidney events preceding death, which was defined as: (i) acute kidney injury as per the Kidney Disease Improving Global Outcomes (KDIGO) stage 3 criteria (creatinine increase of 3.0 times, creatinine increase>353.6 mmol/L, or initiation of acute kidney replacement therapy), or (ii) end-stage kidney disease (defined as chronic kidney replacement therapy, sustained eGFR<15 mL/min/1.73 m^2^, (two eGFR measurements taken at least 28 days apart), or kidney transplantation). The eGFR measurements used for outcome were obtained from one year before death up to the time of death to assess kidney function during the study period.

### Variables

Comorbidities were identified by examining registered ICD-10 codes up to ten years before inclusion (Supplemental Table 1). Concomitant medication at baseline was defined as having redeemed at least one prescription within 180 days before inclusion, based on ATC codes (Supplemental Table 1). Frailty was assessed using the Hospital Frailty Risk Score, a validated and well-described method for categorizing frailty risk^19,20,21^. It utilized a lookback period of 10 years and relied on ICD-10 diagnosis codes obtained from administrative health registers (Supplemental Table 2). Plasma levels of potassium, sodium, and hemoglobin were measured within 90 days before inclusion. Anemia was defined according to the criteria set by the World Health Organization (WHO) as having a plasma hemoglobin level below <7.7 mmol/l (13 g/dl) in males and <7.4 mmol/l (12 g/dl) in females^22^. Hyponatremia was defined as having a plasma sodium level below 135 mmol/l^23^.

### Statistics

Descriptive data at baseline was presented as numbers with percentages for categorical variables, and for continuous variables as medians with interquartile ranges (IQR) or means with standard deviation (SD), as appropriate. Differences across the three eGFR groups by means were compared using the Pearson *χ*^2^ and Kruskal-Wallis tests for categorical and continuous variables, respectively. To calculate the proportion of patients with adverse kidney events preceding death, we used the following formula: Proportion (%) = (Number of patients with one or more adverse kidney events preceding death/ Total number of patients in the cohort) x 100 %. We used linear regression models to calculate eGFR slopes in order to determine the trajectory of eGFR over the past year. Additionally, we conducted subgroup analyses to explore potential factors associated with an increased proportion of patients with adverse kidney events preceding death using chi-square test to test differences in study outcomes. Lastly, for patients who were receiving HF therapy one year prior to death, withdrawal of HF therapy was defined as a 90-day break, and withdrawal rates were calculated across all three eGFR groups up to three months before death.

Two-sided p-values <0.05 were considered significant. All data management and analyses were conducted with SAS, version 9.4 (SAS institute, Cary, NC, USA).

### Ethics

Retrospective register-based studies do not need ethical approval in Denmark. The Danish Data Protection Agency has approved the data access. The study was supported by an unrestricted hospital grant.

## Results

Out of 437,355 deaths in 2014-2021, a total of 50,199 had a previous HF diagnosis. After applying the exclusion criteria, a total of 36,435 patients were included in the final study cohort (Figure 2). A total of 4 % had missing information on creatinine at baseline.

**Figure 2:**
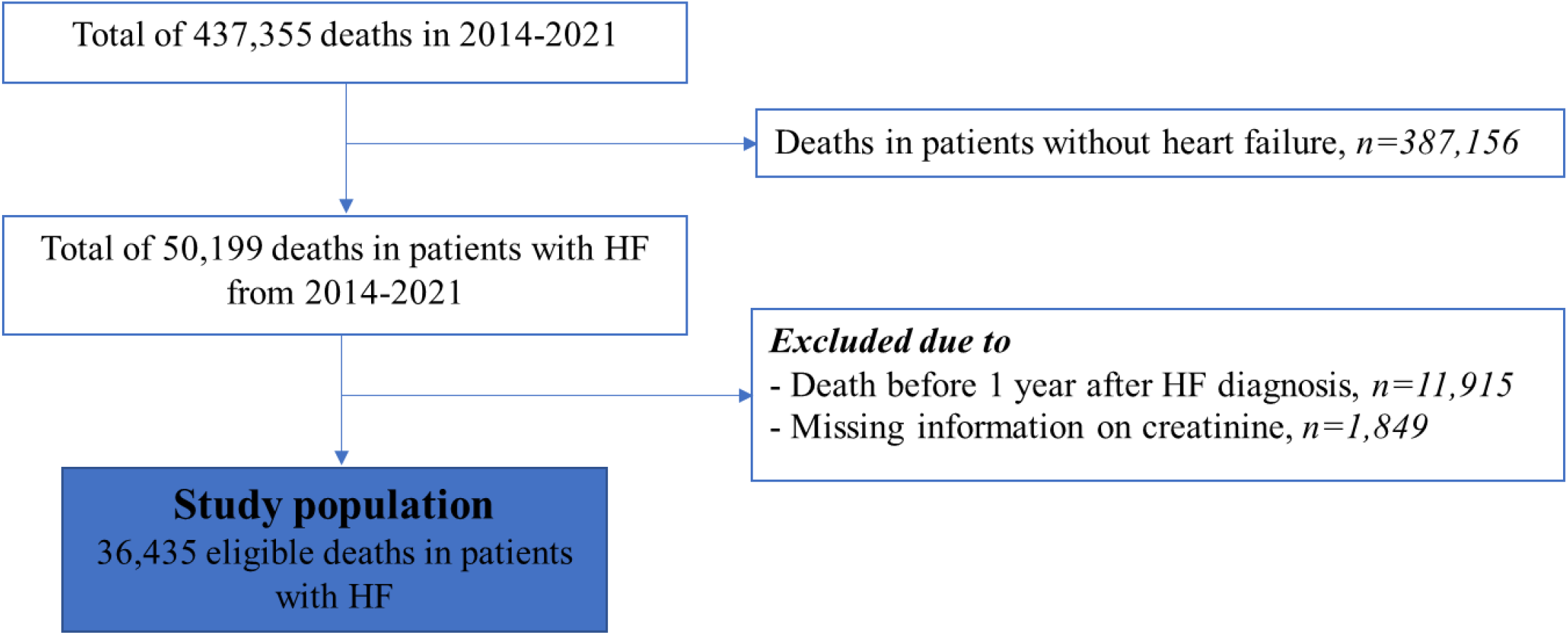
Flowchart

The study population’s characteristics one year prior to death are presented in Table 1. A total of 13,307 (37%) had an eGFR≥60 mL/min/1.73m^2^, 16,756 (46%) had an eGFR between 30 and 59, and 6,372 (17%) had an eGFR<30. The distribution of eGFR at this baseline (one year prior to death) is depicted in Supplemental Figure 1. The median age was 81.1 years (SD 10.6 years), and 61.2 % were men. The median time between obtaining eGFR and the baseline was 27 days (SD 88 days), and the duration of HF was 4.9 years (SD 5.3 years). Among patients with HF who had a preexisting kidney dysfunction, a higher proportion were older, female, were frailer and experienced death in-hospital. The distribution of concomitant pharmacotherapy and comorbidities are illustrated in Table 1.

**Table 1:** Patient characteristics of the study population, 1 year before death stratified by eGFR categories

| | eGFR≥60 | eGFR 30-59 | eGFR<30 |
| --- | --- | --- | --- |
| **Individuals, No. (%)** | 13307 (37) | 16756 (46) | 6372 (17) |
| **Age, No. (%)** |  |  |  |
| <65 years | 2153 (16) | 687 (4) | 363 (6) |
| 65≤age<80 years | 6075 (46) | 5338 (33) | 2026 (32) |
| ≥80 years | 5079 (38) | 10731 (64) | 3983 (63) |
| **Male sex, No. (%)** | 8681 (65) | 9920 (59) | 3693 (58) |
| **Duration of HF in years, mean (SD)** | 4.9 (5.4) | 4.9 (5.3) | 4.8 (5.3) |
| **In-hospital death, No. (%)** | 6402 (48) | 8039 (48) | 3301 (52) |
| **Mean eGFR (SD)** | 79 (15) | 45 (8) | 21 (7) |
| **Plasma potassium, median [IQR]** | 4.0 [3.9, 4.3] | 4.1 [4.0, 4.5] | 4.1 [4.0, 4.6] |
| **Frailty level, No. (%)** |  |  |  |
| Low | 8236 (62) | 9651 (58) | 2909 (46) |
| Intermediate | 3871 (29) | 5545 (33) | 2582 (41) |
| High | 1200 (9) | 1560 (9) | 881 (14) |
| **Nursing home, No. (%)** | 1652 (12) | 2397 (14) | 863 (14) |
| **Comorbidity, No. (%)** |  |  |  |
| Ischemic heart disease | 6059 (46) | 8732 (52) | 3500 (55) |
| Peripheral arterial disease | 1405 (11) | 1919 (12) | 941 (15) |
| Atrial fibrillation | 5510 (41) | 8450 (50) | 3107 (49) |
| Anemia* | 2418 (41) | 2697 (50) | 2063 (66) |
| Hyponatremia** | 854 (14) | 730 (9) | 340 (10) |
| **Treatment, No. (%)** |  |  |  |
| Beta-blocker | 8484 (64) | 11541 (69) | 4602 (72) |
| ACEi or ARB or ARNi | 8354 (63) | 10512 (63) | 3431 (54) |
| Mineralocorticoid receptor antagonist | 3329 (25) | 4387 (26) | 1102 (17) |
| SGLT2 inhibitor | 174 (1) | 136 (0.8) | 19 (0.3) |
| Loop diuretics | 7097 (53) | 12764 (76) | 5353 (84) |
| Thiazide diuretics | 846 (6) | 1090 (7) | 276 (4) |
| Diabetes medication | 2969 (22) | 4217 (25) | 1965 (31) |
| Anticoagulants | 5433 (41) | 8175 (49) | 2636 (41) |
| Antiplatelets | 5611 (42) | 6981 (42) | 2849 (45) |
| Device (ICD/CRT) | 595 (5) | 1038 (6) | 392 (6) |

Values are No (%), mean (SD) or median (IQR). eGFR=estimated glomerular filtration rate; ACEi=angiotensin-converting enzyme inhibitor; ARB=angiotensin receptor blocker; ARNI= angiotensin receptor-neprilysin inhibitor; SGLT2=sodium glucose cotransporter 2; ICD=implantable cardioverter-defibrillator; CRT=cardiac resynchronization therapy. \*Anemia defined as having a plasma hemoglobin level below <7.7 mmol/l (13 g/dl) in males and <7.4 mmol/l (12 g/dl) in females. \*\*Hyponatremia defined as having a plasma sodium level below 135 mmol/l.

A total of 4,774 patients with HF (13.1 %) experienced at least one adverse kidney event within the year preceding their death. AKI was the most frequent, occurring in 9.4 % of patients, while ESKD occurred in 7.7 % of patients during the last year of life. When stratifying based on kidney function one year prior to death, the proportion of patients experiencing AKI was inversely related to kidney function, and this occurred in 6.5 % of patients with eGFR≥60 (95% CI 6.1-6.9), 7.0% of those with eGFR 30-59 (95% CI 6.6-7.4), and 21.9% ((95% CI 20.9-22.9) of those with eGFR<30, p-value for trend <0.001 (Figure 3). The proportions of patients with ESKD were 0.7 % (95% CI 0.6-0.9) for those with eGFR≥60, 2.6% (95% CI 2.4-2.8) for those with eGFR 30-59, and 35.5% (95% CI 34.3- 36.7) for those with eGFR<30, p-value for trend <0.001. The components of AKI and ESKD stratified according to eGFR categories are shown in Supplemental Table 3. Among 2633 patients with a sustained eGFR<15 ml/min/1.73m^2^, 38.0 % did receive kidney replacement therapy in their last year of life (21.4% of those with baseline eGFR≥60, 13.6 % of those with eGFR between 30 and 59, and 42.7% of those with an eGFR<30).

**Figure 3:**
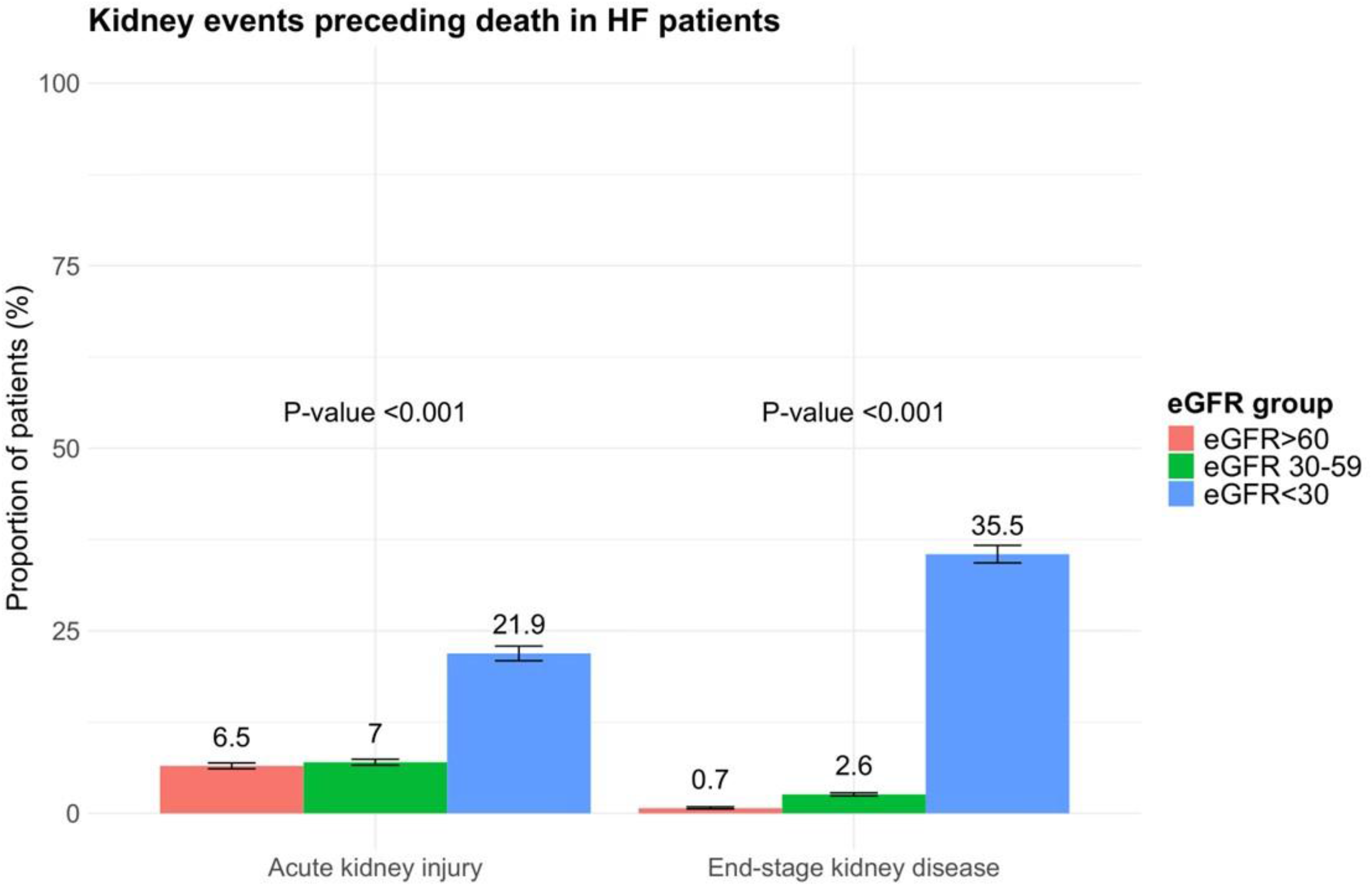
Adverse kidney events preceding death in patients with HF stratified according to eGFR 1 year before death

Figure 4 presents the temporal relationship of the burden of adverse kidney events in the months preceding death, stratified by eGFR. The data show a significant rise in adverse kidney events, including both AKI and ESKD, among patients with HF as they approach the end of life.

**Figure 4:**
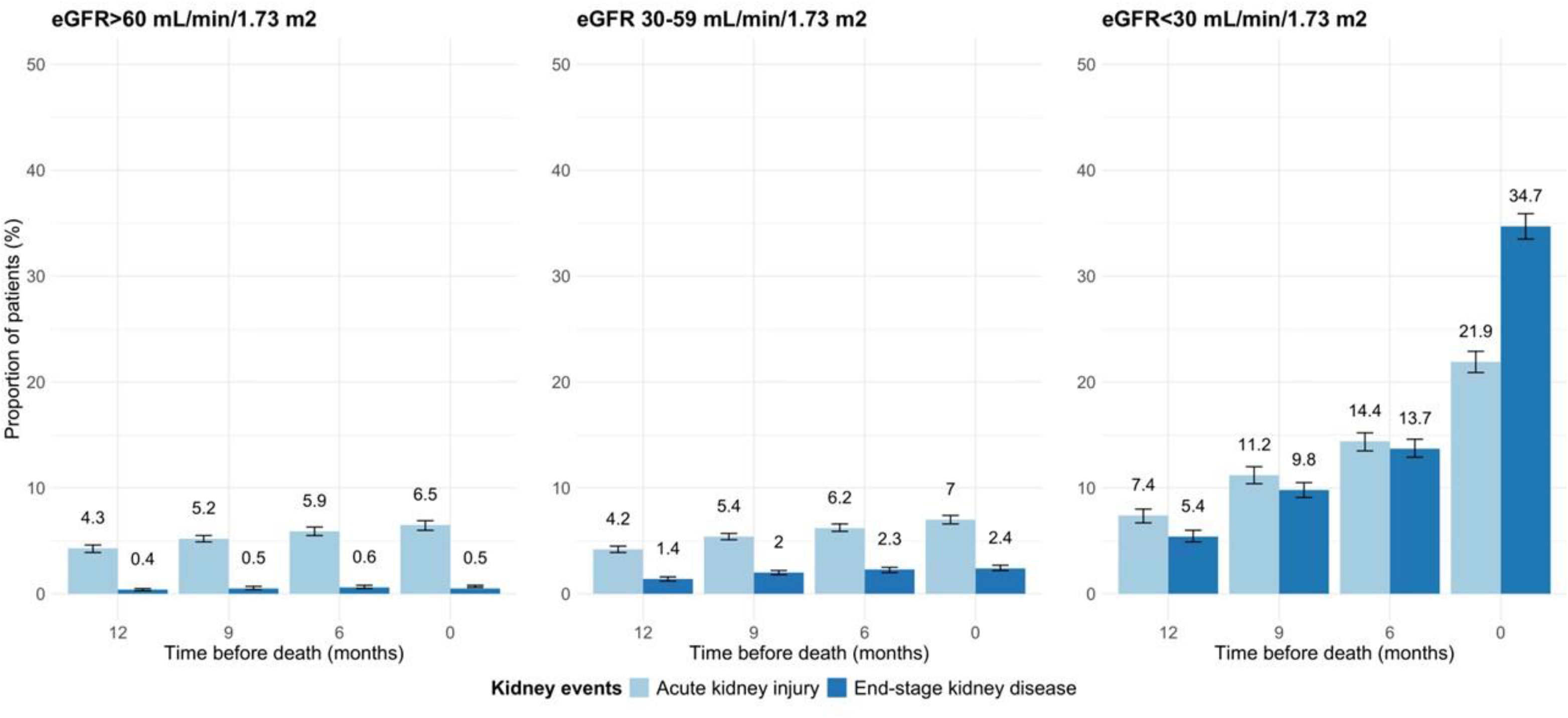
Adverse kidney events preceding death by time before death stratified by eGFR groups 1 year before death

Figure 5 illustrates kidney function trajectories during the end-of-life period, stratified by eGFR groups one year prior to death. Figure 5a depicts the mean eGFR across the three groups. In the final three months before death, there is a significant decline in kidney function, as indicated by decreasing eGFR, particularly among patients with eGFR≥60, whereas eGFR remains stable for patients with eGFR<30 one year prior to death. The decline in eGFR is steepest in patients with an eGFR≥60 during the last 3 months before death (-2.7 mL/min/1.73 m² per month), compared to the period from 12 to 3 months before death (-0.65 mL/min/1.73 m² per month). A similar trend is observed in patients with an eGFR between 30 and 59, with a decline of -1.2 mL/min/1.73 m² per month in the last 3 months, compared to -0.26 mL/min/1.73 m² per month in the 12 to 3 months prior. For patients with an eGFR<30, the change is minimal, with slopes of -0.41 mL/min/1.73 m² per month and -0.08 mL/min/1.73 m² per month, respectively. Figure 5b presents the incidence of chronic kidney replacement therapy initiation among the three groups, demonstrating an increased use of chronic kidney replacement therapy in the last three months.

**Figure 5:**
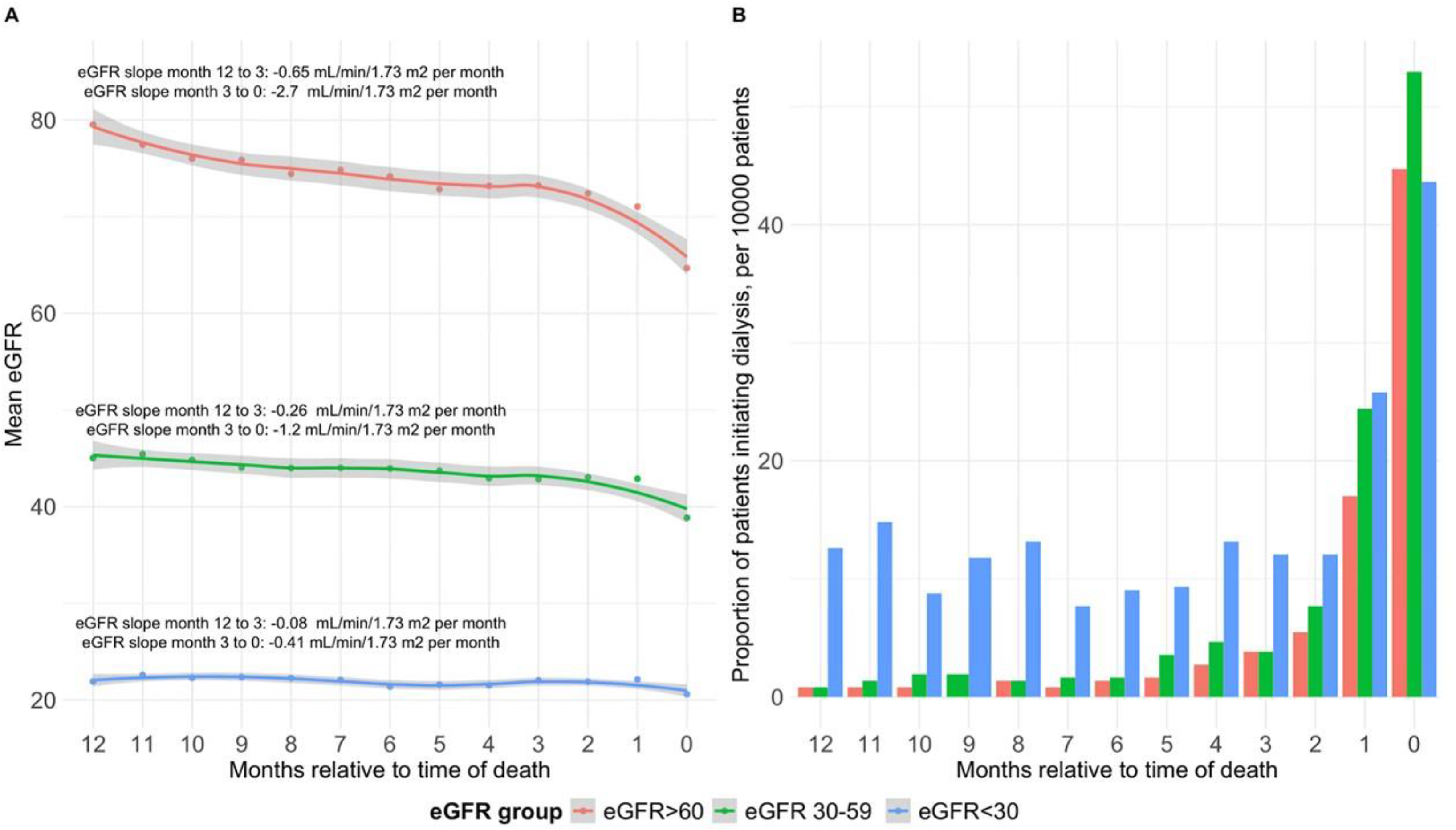
Kidney trajectories during end-of-life stratified by eGFR groups 1 year before death

Subgroup analyses revealed a higher incidence of adverse kidney events including AKI and ESKD preceding death in patients who were male, frailer, and who died at younger age died in-hospital (Table 2). The most pronounced differences were observed between patients who died in-hospital versus those who died out-of-hospital (proportion of patients with AKI 13.9 % vs. 5.2%, proportion of patients with ESKD 9.8% vs 5.7% and between those aged <65 versus those aged ≥80 (18.9 % vs. 5.7 % for AKI and 11.0% vs 5.9 % for ESKD). These findings remained consistent across all components of AKI and ESKD, except for sustained eGFR<15, where the proportion of patients with sustained eGFR<15 was similar across all three age groups. Among patients receiving HF therapy one year before death, withdrawal rates were similar across all three eGFR groups up to 3 months before death (see Supplemental Figure 2).

**Table 2:** Distribution of proportion of patients with any adverse kidney event preceding death stratified by baseline characteristics

|  | **Proportion of AKI**<br>**preceding death** | **P-value** | **Proportion of ESKD**<br>**preceding death** | **P-value** |
| --- | --- | --- | --- | --- |
| **Age group, years** |  |  |  |  |
| **Age<65** | 18.9 % | <0.001 | 11.0 % | <0.001 |
| **65≤age<80** | 12.7 % |  | 9.5 % |  |
| **Age≥80** | 5.7 % |  | 5.9 % |  |
| **Sex** |  |  |  |  |
| **Male** | 10.7 % | <0.001 | 8.2 % | <0.001 |
| **Female** | 7.4 % |  | 6.9 % |  |
| **Place of death** |  |  |  |  |
| **In-hospital death** | 13.9 % | <0.001 | 9.8 % | <0.001 |
| **Out-of-hospital death** | 5.2 % |  | 5.7 % |  |
| **Frailty risk group** |  |  |  |  |
| **Low** | 8.2 % | <0.001 | 5.0 % | <0.001 |
| **Intermediate** | 10.6 % |  | 10.5 % |  |
| **High** | 12.6 % |  | 13.6 % |  |

## Discussion

In this study, we investigated the relationship between kidney function and adverse kidney events leading up to death in patients with HF. Our findings showed that over 1 in 8 HF patients experienced an adverse kidney event in the year before death. AKI was the most common, affecting around 10% of patients, while ESKD occurred in approximately 8% during their final year. Notably, patients with preexisting severely low eGFR had a higher incidence of adverse kidney events, particularly in the last three months of life.

Emerging evidence shows that SGLT-2 inhibitors, the steroidal MRA finerenone, and sacubitril/valsartan positively impact both kidney and cardiac outcomes ^7,24,25,26,27,28^ across a range of kidney function levels ^29–31^. These drugs have proven to be both effective and safe for patients with varying degrees of kidney impairment. Despite the importance of consistently assessing and reporting kidney outcomes in HF trials, traditional HF trials often fall short in adequately capturing kidney-related outcomes, particularly kidney-related mortality ^32^. While newer HF trials have incorporated kidney safety endpoints, they are not primarily powered to analyze kidney-specific outcomes, few patients progress to dialysis during the trial period, and those with advanced kidney disease are frequently excluded.

Our findings of a high incidence of adverse kidney events in the final year of life among HF patients suggest that kidney-related mortality may be underrecognized and underreported in clinical trials. While we observed a temporal association between adverse kidney events and death, causality cannot be established, nor can we definitively attribute these deaths to kidney-related causes. However, the true incidence of kidney-related deaths is likely higher than reported, particularly given the limitations in current definitions. Defining kidney-related death in the context of HF and chronic kidney disease (CKD) is complex. Clinical manifestations such as volume overload, dyspnea, and elevated natriuretic peptide levels can result from CKD progression, worsening HF, or a combination of both. Existing trial definitions, which often classify kidney-related death as death due to ESKD without initiation of kidney replacement therapy, may fail to capture the full spectrum of kidney-related mortality ^11^. For instance, only one case of kidney death was reported in the DAPA-HF trial ^9^, and two cases in the PARAGON-HF trial ^33^. Similarly, kidney-related deaths were minimally reported in CKD trials ^34^. This highlights the need for more comprehensive reporting and better characterization of kidney-related outcomes in both HF and CKD trials. Such efforts will improve the accuracy of mortality assessments, especially since worsening kidney function may reflect the progression of HF rather than a direct cause of death. Moreover, other factors, such as sepsis, may contribute to both declining kidney function and worsening HF, further complicating the attribution of kidney-related mortality. This reinforces the need for novel therapies aimed at preventing kidney function decline and more precise definitions of kidney-related death in clinical research.

Our study revealed a high incidence of adverse kidney events in patients with preexisting eGFR <30, aligning with pooled analyses from the EMPEROR-Preserved and EMPEROR-Reduced trials, where patients at higher KDIGO risk faced an increased likelihood of experiencing a composite kidney event^30^. Conversely, in the PARADIGM-HF trial, a composite kidney outcome consisting of ESKD and sustained eGFR decline of <40% occurred equally across the three KDIGO risk groups^29^. Patients with preexisting eGFR<30 appeared more vulnerable to kidney injury, with a higher incidence of comorbidities and greater use of loop diuretics, suggesting limited renal reserve and a greater likelihood of progressing to dialysis, compared to the patients with higher eGFR, which more commonly used ACEi/ARB/SGLT2i/MRA. The high incidence of acute kidney injury episodes in the last month before death suggests that deteriorating kidney function in the late stages of HF may contribute to the fatal trajectory of the disease. However, it is also possible that the worsening of HF itself leads to declining kidney function.

Another surrogate endpoint for kidney failure is the eGFR slope which has been shown to be a valid, appropriate, and reliable surrogate marker for kidney failure^35^, especially in situations where trials may lack sufficient power to evaluate major renal outcomes^36^. In this study, the decline in eGFR was most highly significant for patients with an eGFR≥60 and during the last three months before death, with a slope reaching as steep as -2.7 mL/min/1.73 m² per month in the last three months, which was > 4 times the decline in the preceding 9 months and corresponding to a slope of -32.4 mL/min/1.73 m^2^ per year. The higher baseline eGFR of patients in this group may partly explain the greater decline in eGFR observed, a phenomenon attributable to regression to the mean. In PARADIGM-HF, patients in the lowest KDIGO risk category had the highest annual decline in eGFR with slope of -1.7 mL/min/1.73 m² per year^29^. This was consistent with DAPA-HF and DELIVER with slope of -2.3 mL/min/1.73 m² per year in the placebo group^37^.

Our subgroup analyses provide valuable insights into the factors associated with a higher proportion of patients experiencing adverse kidney events. The proportion of adverse kidney events was higher for patients who died at a younger age compared to those aged 80 years and above. This finding was consistent across all components of the composite endpoints AKI and ESKD, except for sustained eGFR<15, where the distribution was equal among all three age groups. This disparity may be due to clinicians being more likely to initiate kidney replacement therapy in younger patients, however the kidney burden appears to be higher for younger patients, whereas older patients may have other contributing factors for their death, such as sepsis and infections. Additionally, our analyses revealed that male sex was associated with a higher proportion of adverse kidney events preceding death in patients with HF. This finding aligns with previous studies suggesting that male sex may be a risk factor for kidney dysfunction in cardiovascular diseases^38^. Patients who died in-hospital had a higher proportion of any adverse kidney events, and this underscores the importance of considering the critical role of acute decompensations and hospitalizations in the progression of kidney dysfunction. Acute kidney injury during hospitalizations for HF exacerbations may exacerbate the underlying kidney impairment and contribute to adverse outcomes. The association between higher degrees of frailty and adverse kidney events preceding death in patients with HF highlights the significance of considering the overall health status of patients in HF management.

### Clinical perspectives

The high frequency of adverse kidney events in patients with HF underscores the importance of monitoring and managing kidney function in this vulnerable population. To enhance patient care, future research should focus on exploring the underlying mechanisms linking HF and kidney dysfunction, identifying risk factors for kidney death, and developing targeted interventions to mitigate the impact of kidney complications on HF prognosis.

### Strengths and Limitations

The primary strength of this study lies in its comprehensive inclusion of all deaths among patients diagnosed with HF in Denmark, utilizing national registries. However, it is crucial to acknowledge certain limitations inherent in the study design, which restricts the ability to establish causal inferences based on observed differences in the registry-based data. An important limitation of the study is the lack of available data on certain clinical parameters, including degree of anuria, blood pressure, NT-proBNP, LVEF, albuminuria, cystatin C, cardiac index, pulmonary capillary wedge pressure, and the etiology of kidney dysfunction (such as exposure to nephrotoxins like contrast dye or nonsteroidal anti-inflammatory drugs or hemodynamic deterioration). The absence of this information may have resulted in residual and unmeasured confounding, potentially impacting the strength of associations observed in the study. Additionally, a minimum HF duration of 12 months before death was required, thus excluding patients with fulminant HF who had less than 12 months of follow-up. The impact of medication changes on eGFR evolution during follow-up was not analyzed, presenting a potential limitation. Despite these limitations, the study provides valuable insights into the occurrence of adverse kidney events preceding death in patients with HF, serving as a basis for further research and clinical consideration in this critical area.

## Conclusion

In this real-world study, AKI and ESKD are common in the last year of life among patients with HF, particularly in those with a pre-existing lower eGFR and particularly in the last months before death. Further research and awareness of kidney complications are necessary to improve outcomes in this population, and a definition of the cause of death due to kidney complications is needed. With the present definition of kidney death or lack thereof kidney death may be underestimated in HF patients.

## Abbreviations list

CKD: Chronic kidney disease AKI: Acute kidney injury ESKD: End-stage kidney disease

eGFR: Estimated glomerular filtration rate HF: Heart failure

ICD-10: 10th edition of the International Classification of Diseases KDIGO: Kidney Disease Improving Global Outcomes

LVEF: Left ventricular ejection fraction

## Data Availability

The data that support the findings of this study are derived from Danish administrative registries, which are not publicly available due to legal and privacy restrictions under Danish data protection regulations. Access to these data can only be granted through secure research environments managed by Danish authorities. Researchers may request access by contacting the corresponding author, subject to approval from the relevant data protection agency and in accordance with institutional and ethical guidelines.

## Figure legends

Figure 1: Study timeline

All deaths in 2014-2021 that occurred in patients after at least 1 year with HF. Baseline was defined as 1 year prior to death. The exposure assessment period correspondingly extended up to 1 year prior to death, except for kidney transplantation, which had a 15-year lookback period.

Figure 2: Flowchart

Flowchart of the study population with exclusion criteriums.

Figure 3: Adverse kidney events preceding death in patients with HF stratified according to eGFR 1 year before death

Proportion of adverse kidney events in the three eGFR groups up to one year before time of death.

Figure 4: Adverse kidney events preceding death by time before death stratified by eGFR groups 1 year before death

Proportion of adverse kidney events in the three eGFR groups up to one year before time of death stratified in time before death

Figure 5: Kidney trajectories during end-of-life stratified by eGFR groups 1 year before death

Figure 5a: Mean eGFR during last year before death.

Figure 5b: Initiation of kidney replacement therapy during last year before death.

